# Is *GBA1* T369M not a risk factor for Parkinson’s disease in the Swedish population?

**DOI:** 10.1101/2024.03.15.24304347

**Authors:** Kajsa Atterling Brolin, David Bäckström, Joel Wallenius, Ziv Gan-Or, Andreas Puschmann, Oskar Hansson, Maria Swanberg

## Abstract

Variants in *GBA1* are important genetic risk factors in Parkinson’s disease (PD). *GBA1* T369M has been linked to an ∼80% increased PD risk but the reports are conflicting and the relevance of *GBA1* variants in different populations varies. A lack of association between T369M and PD in the Swedish population was recently reported but needs further validation. We therefore investigated T369M in 1,808 PD patients and 2,183 controls and our results support that T369M is not a risk factor for PD in the Swedish population.

## Introduction

Multiple loci have been identified to be linked to Parkinson’s disease (PD), of which *GBA1* is one of the most important [1, 2]. *GBA1* encodes the lysosomal enzyme glucocerebrosidase and biallelic pathogenic variants cause the autosomal recessive storage disorder Gaucher disease (GD) [3]. Notably, *GBA1* is a pleomorphic locus with both coding, structural, and non-coding variants classified as mild or severe depending on the type of GD they cause [2, 4, 5]. Patients with GD and heterozygous carriers of pathogenic *GBA1*-variants have an increased PD risk and *GBA1* variants also affect PD age at onset (AAO) [2, 3]. Additional variants that are not causal for GD have been reported to be risk factors for PD, including *GBA1* T369M (c.1223C>T p.[Thr408Met], rs75548401, chr1:155206037G>A [GRCh37]) [1].

*GBA1* T369M was linked to increased PD risk in a meta-analysis, with an odds ratio (OR) of 1.78 (95% confidence interval [CI]=1.25–2.53, p=0.002) [6]. Subsequently, some studies have reported a significant association with an OR=1.53 (p=0.0012) [7], OR=2.54 (95% CI=1.30–4.95, p=0.0094) [8] and OR=4.98 (95% CI=1.48-16.66, p=9.25E-03) [9]. Additional studies report a higher T369M frequency in PD patients but a lack of association with PD [10-12] and another report a higher frequency in the control group in an Irish study [13].

Recently, Ran and colleagues reported lack of association between T369M and PD in 1,131 PD patients and 1,594 controls in Sweden [14]. However, the authors report possible problems with the control population, which displayed an unusually high allele frequency and two homozygous AA carriers. The variant was also not in Hardy Weinberg Equilibrium (HWE) in entire control material and a proportion of the control population were blood donors of unknown age, and these individuals could develop PD later in life

In this study, we further investigated whether *GBA1* T369M is a risk factor for PD in Sweden, using separate datasets from two case-control cohorts, one PD cohort with enriched PD family history, and two large population-based cohorts.

## Methods

### Participants and genotyping

The study was approved by the regional ethics review board of Lund (2013/509 and 2006/615), and Umeå (03-387, 2014-163-31M, 2020-04074, 2021-03965, 2022-01219-01) and by the Swedish Ethical Review Authority (2022-03840-01 and 2019-05036). All participants gave informed written consent at study enrollment in each of the datasets.

A dataset of 1,808 PD patients and 2,183 controls from two sites in Sweden, Lund (MultiPark biobank sample collection [MPBC]) and Umeå, was analyzed. The dataset from Lund has been described in detail previously [15] . In brief: PD patients (ICD-10 G20) and population-based controls matched on year of birth, sex and residential area were genotyped with the Infinium Global Screening Array-24 v.2.0 (GSAMD-24v2.0). The Umeå site had previously included individuals with a possible PD diagnosis (ICD-10 G20, G21 and G23) by which a PD diagnosis was validated through neurological medical records by a specialist in neurology and movement disorders. For most PD patients (>80%), samples had been donated to the neurobiobank of the neurology clinic at the University Hospital of Umeå. Remaining PD cases were identified in the Northern Sweden Health and Disease Cohort (NSHDC), containing samples from >135,000 Västerbotten individuals, and controls were matched by age, sex, and residential area. Genotype data using the GSA v1.0 had previously been obtained from deCODE Genetics/Amgen.

The datasets SweGen and the EpiHealth Elderly cohort (EpiHealth), were used to determine the frequency in the general Swedish population. Data from SweGen is available through SweFreq — The Swedish Frequency resource for genomics (https://swefreq.nbis.se/) and was obtained 29 February 2024. It contains whole-genome variant frequencies for 1,000 Swedes, representing the genetic variation in the population with a median age at sampling of 65 years [16]. EpiHealth is a population-based cohort of ∼25,000 individuals genotyped with the GSAMD-24v2.0 and imputed with the Haplotype reference consortium panel [17]. Individuals <71 years, closely related individuals, individuals of non-European ancestry and individuals with a self-reported PD diagnosis were removed prior to analysis, resulting in 3,049 individuals.

We additionally investigated a dataset with whole exome sequencing data from 158 well-characterized PD patients from southern Sweden of whom 61.7% reported a first or second degree relative with PD (MultiPark NGS database, https://www.multipark.lu.se/infrastructures/ngs-database-infrastructure.)

### Statistical analyses

Power calculations were done using the Genetic Association Study (GAS) Power Calculator (http://csg.sph.umich.edu/abecasis/gas_power_calculator/). At a significant level p=0.05, considering a sample size of 1,808 PD patients and 2,183 controls, a disease prevalence of 0.2%, and an estimated minor allele frequency (MAF) of 1-2% (gnomAD Euro/Finnish), we have 80% power at OR=1.51-1.75. The lowest OR reported for *GBA1* T369M on the *GBA1*-PD browser is 1.4 (Parlar 2023, PMID: 36598340), which we with the current sample size have 62% and 36% power to identify at a MAF of 2% and 1%, respectively.

MAFs, associations and HWE for T369M were calculated using PLINK v1.9 [18, 19]. HWE was calculated for all cohorts separately as well as the combined dataset and for both the patient and control group within the cohorts. For the SweGen dataset and the smaller PD dataset, HWE was calculated using the Michael H. Court online HWE calculator (https://accounts.smccd.edu/case/biol215/docs/HW_calculator.xls). Genetic analysis of genotypes and alleles was performed using Fisher’s exact tests in PLINK v1.9.

## Results

The frequency of *GBA1* T369M was first investigated in a dataset of 1,808 PD patients and 2,183 controls (Table 1), in which no association with PD was observed (OR=1.14, 95% CI=0.78–1.68, p=0.491). The MAF was 1.44% in patients and1.26% in controls. The heterozygous GA genotype frequency was 2.88% for patients and 2.53% for controls. No homozygous AA carriers were identified. The variant was in HWE in both the separate and combined datasets (p=1.00). As *GBA1* variants can affect PD AAO, we further investigated the mean AAO in the different genotype groups. For the MPBC dataset, AAO was not available and age at diagnosis (AAD) was used as a proxy for AAO. We observed an identical AAO of ∼64 years for both the homozygous GG carriers and heterozygous carriers in the combined dataset. No difference in AAO was observed in the separate datasets with ∼63 (GG homozygous) vs ∼64 years (heterozygous) for the Umeå dataset and ∼65 years in the MPBC dataset for both groups. Among individuals with early onset PD (EOPD), defined as diagnosis at the age of <50 years according to the Movement Disorder Society Task Force on EOPD [20], only two heterozygous carriers (1.29%) were observed. The sex distribution was also similar for the homozygous and heterozygous carriers with ∼38% and 37% women, respectively.

**Table 1:**
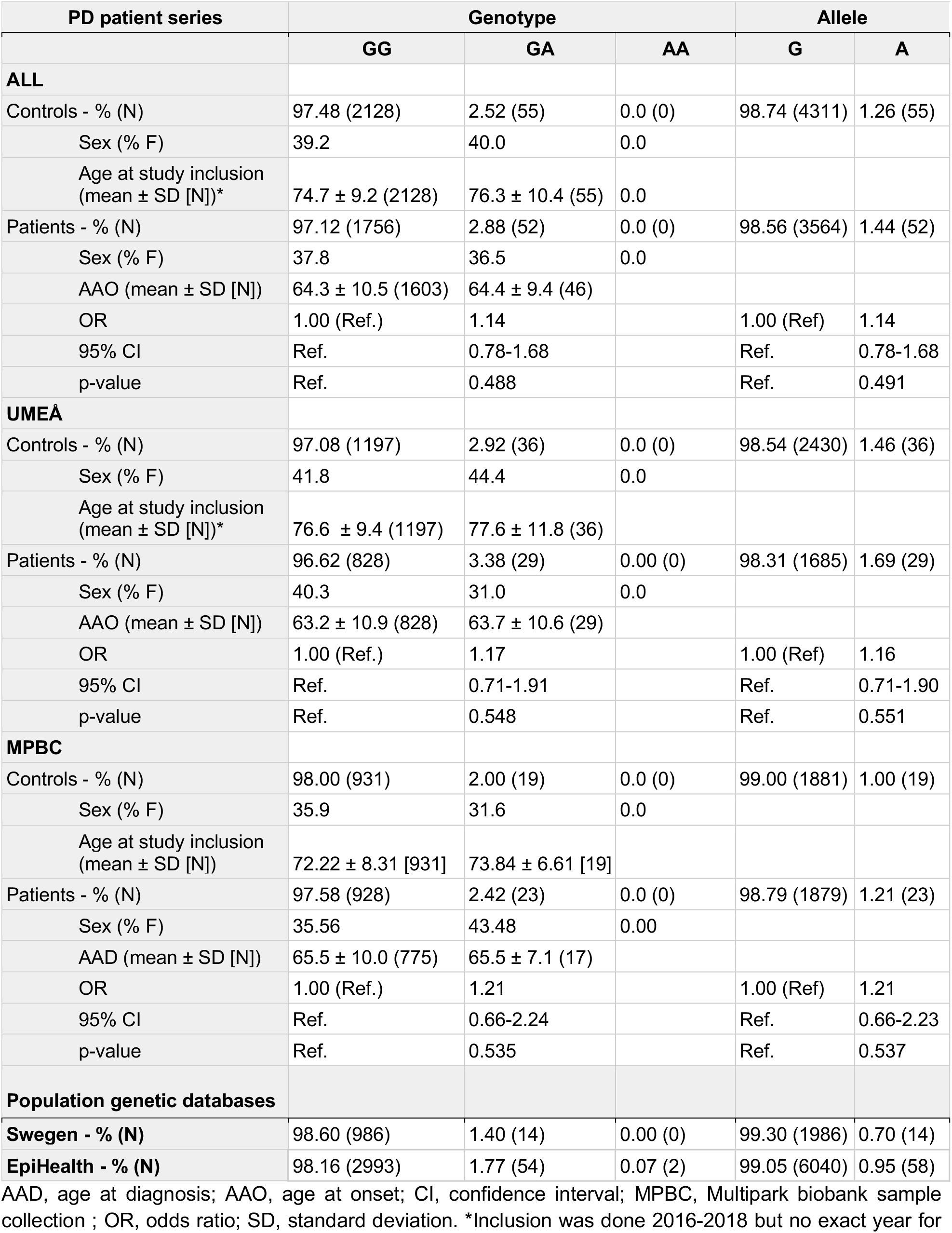
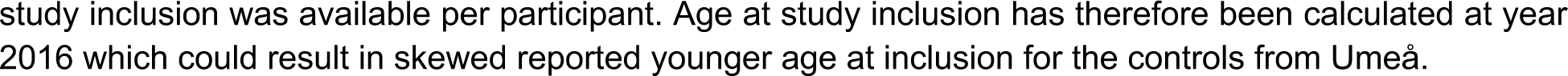
Genotype and allele frequencies of *GBA1* T369M in Sweden.

| PD patient series | Genotype |  |  | Allele |  |
| --- | --- | --- | --- | --- | --- |
|  | GG | GA | AA | G | A |
| **ALL** |  |  |  |  |  |
| Controls - % (N) | 97.48 (2128) | 2.52 (55) | 0.0 (0) | 98.74 (4311) | 1.26 (55) |
| Sex (% F) | 39.2 | 40.0 | 0.0 |  |  |
| Age at study inclusion (mean ± SD [N])* | 74.7 ± 9.2 (2128) | 76.3 ± 10.4 (55) | 0.0 | | |
| Patients - % (N) | 97.12 (1756) | 2.88 (52) | 0.0 (0) | 98.56 (3564) | 1.44 (52) |
| Sex (% F) | 37.8 | 36.5 | 0.0 |  |  |
| AAO (mean ± SD [N]) | 64.3 ± 10.5 (1603) | 64.4 ± 9.4 (46) | | | |
| OR | 1.00 (Ref.) | 1.14 |  | 1.00 (Ref) | 1.14 |
| 95% CI | Ref. | 0.78-1.68 |  | Ref. | 0.78-1.68 |
| p-value | Ref. | 0.488 |  | Ref. | 0.491 |
| **UMEA** |  |  |  |  |  |
| Controls - % (N) | 97.08 (1197) | 2.92 (36) | 0.0 (0) | 98.54 (2430) | 1.46 (36) |
| Sex (% F) | 41.8 | 44.4 | 0.0 |  |  |
| Age at study inclusion (mean ± SD [N])* | 76.6 ± 9.4 (1197) | 77.6 ± 11.8 (36) | | | |
| Patients - % (N) | 96.62 (828) | 3.38 (29) | 0.00 (0) | 98.31 (1685) | 1.69 (29) |
| Sex (% F) | 40.3 | 31.0 | 0.0 |  |  |
| AAO (mean ± SD [N]) | 63.2 ± 10.9 (828) | 63.7 ± 10.6 (29) | | | |
| OR | 1.00 (Ref.) | 1.17 |  | 1.00 (Ref) | 1.16 |
| 95% CI | Ref. | 0.71-1.91 |  | Ref. | 0.71-1.90 |
| p-value | Ref. | 0.548 |  | Ref. | 0.551 |
| **MPBC** |  |  |  |  |  |
| Controls - % (N) | 98.00 (931) | 2.00 (19) | 0.0 (0) | 99.00 (1881) | 1.00 (19) |
| Sex (% F) | 35.9 | 31.6 | 0.0 |  |  |
| Age at study inclusion (mean ± SD [N]) | 72.22 ± 8.31 [931] | 73.84 ± 6.61 [19] | | | |
| Patients - % (N) | 97.58 (928) | 2.42 (23) | 0.0 (0) | 98.79 (1879) | 1.21 (23) |
| Sex (% F) | 35.56 | 43.48 | 0.00 |  |  |
| AAD (mean ± SD [N]) | 65.5 ± 10.0 (775) | 65.5 ± 7.1 (17) | | | |
| OR | 1.00 (Ref.) | 1.21 |  | 1.00 (Ref) | 1.21 |
| 95% CI | Ref. | 0.66-2.24 |  | Ref. | 0.66-2.23 |
| p-value | Ref. | 0.535 |  | Ref. | 0.537 |
| **Population genetic databases** |  |  |  |  |  |
| **Swegen - % (N)** | 98.60 (986) | 1.40 (14) | 0.00 (0) | 99.30 (1986) | 0.70 (14) |
| **EpiHealth - % (N)** | 98.16 (2993) | 1.77 (54) | 0.07 (2) | 99.05 (6040) | 0.95 (58) |

AAD, age at diagnosis; AAO, age at onset; CI, confidence interval; MPBC, Multipark biobank sample collection ; OR, odds ratio; SD, standard deviation. \*Inclusion was done 2016-2018 but no exact year for
study inclusion was available per participant. Age at study inclusion has therefore been calculated at year 2016 which could result in skewed reported younger age at inclusion for the controls from Umeå.

We further investigated T369M in two population-based datasets, SweGen and EpiHealth, with 1,000 and 3,039 individuals, respectively. The MAF was 0.70% in SweGen and 0.95% in EpiHealth (Table 1). T369M was in HWE in SweGen (p=0.810) but not in EpiHealth (p=0.029). No homozygous AA carriers were identified in SweGen but two were identified in EpiHealth. The variant was imputed in EpiHealth with an rsq=0.819. Twenty-four self-reported PD patients were identified in EpiHealth, all homozygous GG-carriers which were excluded.

The allele frequency of T369M was higher in the dataset of PD patients with enriched PD family history, 2.85% with no homozygous AA carriers and a heterozygous carrier frequency of 5.70% (Table 2). The variant was also in HWE in this dataset (p=0.713). A younger mean AAO of ∼57 years was observed for the heterozygous carriers as compared to the homozygous GG carriers, ∼60 years. However, this could be caused by the few numbers of heterozygous carriers as the median AAO for the groups was 62 vs 61 years, respectively (Table 2).

**Table 2:** Genotype and allele frequencies of *GBA1* T369M in 159 PD patients from the MultiPark NGS database.

|  | Genotype |  |  | Allele |  |
| --- | --- | --- | --- | --- | --- |
|  | GG | GA | AA | G | A |
| **Patients - % (N)** | 94.30 (149) | 5.70 (9) | 0.00 (0) | 97.15 (307) | 2.85 (9) |
| Sex (% F) | 36.2 | 55.6 | 0.0 |  |  |
| AAO (mean ± SD [N]) | 59.6 ± 10.9 (136) | 56.78 ± 14.13 | | | |
| AAO (median; IQR [N]) | 61.0; 13.3 (136) | 62.0; 19.0 (127) |  |  |  |

AAO, age at onset; IQR, interquartile range; SD, standard deviation

## Discussion

In this study we have performed the largest analysis of *GBA1* T369M in PD in Sweden in a separate dataset to that previously used by Ran and colleagues [14]. Our results further support that T369M is not a risk factor for PD in the Swedish population. Nevertheless, it cannot be excluded that it has an effect on PD risk in Sweden as we observed a slightly higher MAF in patients compared to controls (1.44 vs. 1.26%) and even higher frequency in the dataset with enriched PD family history (2.85%). Notably, the two population-based datasets had lower MAFs than the controls in the combined PD case-control dataset (0.70% and 0.95%) which could argue that the MAF in the controls is higher than expected.

However, the importance of a geographically matched control group for genetic analyses in Sweden has previously been emphasized [16, 21] and we consider the matched control groups in our datasets appropriate. All analyses were performed in individuals of European ancestry but no adjustment for potential population stratification was done. Two homozygous AA carriers were identified in one of the population-based datasets, EpiHealth, both females included at the age of 74 with no reports of a PD diagnosis at the time of inclusion. However, we cannot exclude the possibility that these individuals develop PD at an older age. Additionally, the variant was imputed and deviated from HWE in this dataset, which could result in false positive carriers. The result from this dataset should thus be interpreted with caution.

Despite the potential lack of an association, T369M could still contribute to PD risk in Sweden as it has been reported that a second risk variant in *GBA1* is the most important contributor to disease risk [22]. This could explain the higher frequency observed in the dataset with PD patients with enriched PD family history. When investigating other coding variants in the 9 identified heterozygous carriers in this dataset, one individual was compound heterozygous for *GBA1* E326K. Noteworthy, we have not investigated compound heterozygosity in any of the other datasets, and other *GBA1* variants i.e., L444P and E326K are strongly associated with PD in Sweden (OR=8.17, 95%CI=2.51-26.23, p=0.002 and OR=1.60, 95%CI=1.16-2.22, p=0.026) [23].

It is clear that *GBA1* plays a vital role in PD and it became even more apparent following the recent finding of a novel genetic risk factor in *GBA1* in people of African ancestry, which has not been seen in European populations [4]. It is thus of importance to evaluate *GBA1* variants in various populations as their frequency varies from ∼2% to ∼20% [24]. Our results show that the frequency of T369M is low in PD patients in Sweden and support previous findings that *GBA1* T369M is not a risk factor for PD in the Swedish population.

## Data Availability

The data supporting the findings of this study from the individual datasets are available upon reasonable request from the corresponding author or co-authors. The EpiHealth Elderly cohort is available for researchers following application and approval. The data for all datasets are not publicly available due to privacy or ethical restrictions. Frequency data for SweGen are openly available in the SweGen Variant Frequency Dataset at https://swefreq.nbis.se/.

## Acknowledgements

The authors would like to thank the participants who donated their time and biological samples, making this study possible. We would also like to thank the Swedish Research Council for supporting the strategic research network Epidemiology for Health (EpiHealth) and Multidisciplinary research focused on Parkinson’s disease (Multipark) and thereby also the EpiHealth screening cohort and the steering committee of the MultiPark’s biobank sample collection.

